# Explainable Artificial Intelligence to Diagnose Early Parkinson’s Disease via Voice Analysis

**DOI:** 10.1101/2024.09.29.24314580

**Authors:** Matthew Shen, Pouria Mortezaagha, Arya Rahgozar

**Affiliations:** St. Theresa of Lisieux Catholic High School, Toronto; Ottawa Hospital Research Institute, University of Ottawa School of Engineering Design and Teaching Innovation, Ottawa

**Keywords:** Parkinson’s Disease, Deep Learning, Vocal Biomarkers, Explainable AI

## Abstract

**Background:** Parkinson’s disease (PD) is a progressive neurodegenerative disorder that affects motor control, leading to symptoms such as tremors or impaired balance. Early diagnosis of PD is crucial for effective treatment, yet traditional diagnostic models are often costly and lengthy. This study explores the use of Artificial Intelligence (AI) and Machine Learning (ML) techniques, particularly voice analysis, to identify early signs of PD and make a precise diagnosis.

**Objectives:** This paper aims to create an automatic detection and prediction of PD binary classification using vocal biomarkers. We will also use explainability to identify latent and important patterns in the input data in retrospect to the target to inform the definition of Parkinson’s through voice characteristics. Finally, a probability generation will be generated to create a scoring system of a patient’s odds of PD as a spectrum.

**Methods:** We utilized a dataset comprising 81 voice recordings from both healthy control (HC) and PD patients, applying a hybrid AI model combining Convolutional Neural Networks (CNN), Recurrent Neural Networks (RNN), Multiple Kernel Learning (MKL), and Multilayer Perceptron (MLP). The model’s architecture was designed to extract and analyze acoustic features such as Mel-Frequency Cepstral Coefficients (MFCCs), local jitter, and local shimmer, which are all indicative of PD-related voice impairments. Once features are extracted, the AI model will generate prediction labels for HC or PD files. Then, a scoring system will assign a number ranging from 0-1 to each file, indicating the stage of PD development.

**Results:** Our champion model yielded the following results: diagnostic accuracy of 91.11%, recall of 92.50%, precision of 89.84%, an F1 score of 0.9113, and an area under curve (AUC) of 0.9125. Furthermore, the use of SHapley Additive exPlanations (SHAP) provided detailed insight into the model’s decision-making process, highlighting the most influential features contributing to a PD diagnosis. The outcomes of the implemented scoring system demonstrate a distinct separation in the probability assessments for PD across the 81 analyzed audio samples, validating our scoring system by confirming that the vocal biomarkers in the audio files accurately correspond with their assigned scores.

**Conclusion:** This study highlights the efficacy of AI, particularly a hybrid model combining CNN, RNN, MKL, and Deep Learning in diagnosing early PD through voice analysis. The model demonstrated a robust ability to distinguish between HC and PD patients with significant accuracy by leveraging key vocal biomarkers such as MFCCs, jitter, and shimmer.

## 1 Introduction

Parkinson’s disease is a disorder of the central nervous system. It causes unintentional and uncontrollable bodily movements such as shaking, stiffness, or difficulty with balance and control. PD is a neurodegenerative disorder, meaning the symptoms gradually worsen over time. Due to this, people suffering from PD may develop behavioral or mental changes such as depression or a decrease in memory. Currently, there is no cure for PD, but there are medications that can alleviate symptoms. Regardless, it is best to intervene and prevent the gradual onset of PD rather than treating it at its most vicious state. However, traditional diagnostic methods often rely on clinical evaluations and imaging techniques, which can be invasive, costly, and require specialized medical expertise. In recent years, the advent of AI has opened new opportunities for diagnosis, particularly through voice analysis. This paper explores the use of AI and ML techniques to diagnose early-stage PD by analyzing vocal characteristics. It aims to provide a comprehensive review of current methodologies, findings, and future avenues in this rapidly growing field.

Recent advancements in AI and ML have demonstrated significant potential in diagnosing Parkinson’s disease using voice analysis. Various studies have utilized the extracted acoustic features of voice recordings to distinguish between healthy individuals and individuals with PD. While traditional statistical methods have been employed, the field is rapidly evolving towards the use of deep learning techniques that automatically extract relevant features from raw voice data.

### 1.1 Voice analysis techniques

Little et al. used support vector machines (SVM) to classify voice recordings of PD patients with an accuracy of 91.4%, establishing themselves as one of the first pioneers in this field [1]. Their study demonstrated the viability of using acoustic voice features for PD diagnosis and laid the groundwork for further research. However, this study lacked MFCCs, which are instrumental for projects using voice to diagnose PD. This paper will incorporate MFCCs alongside traditional acoustic features to ensure a thorough diagnosis. Building on this, Tsanas et al. developed a decision support system using MKL to replicate the unified Parkinson’s disease rating scale (which requires the patient’s presence in the clinic) remotely [2]. Their approach underscored the importance of integrating multiple learning features and robust ML techniques when transitioning to noninvasive and self-administered PD tests. More recent studies, however, focus on deep learning models—automatic extraction of relevant features from raw voice data. For example, Alhanai et al. employed a Long-Short Term Memory (LSTM) neural network to analyze speech patterns with an 89% accuracy in detecting early PD symptoms [3]. Similarly, Alissa et al. used a CNN to extract and analyze voice features, achieving a diagnosis accuracy of 93.5% [4]. These studies highlight a transition from traditional methods to more sophisticated AI models.

### 1.2 Multimodal approaches

Integrating voice analysis with other modalities, such as data from wearable devices, has shown promise in improving diagnostic accuracy. For example, Guo et al. demonstrated that combining voice data with other physiological signals improved their overall accuracy of PD diagnosis to around 96.06% [5]. However, this area of study is relatively novel, and researchers are still experimenting with ways to accurately combine voice and locomotive movement. Our paper only uses voice as data input, isolating the model’s accuracy so it disregards any other biomarker. Removing confounding variables lets us properly gauge how important vocal biomarkers are for diagnosing PD.

### 1.3 Model architecture

Historically, AI model applications in medical analysis have used decoupled model architectures. This means the model does not leverage multiple networks concurrently. A notable exception in recent literature is a pipeline AI model that uses SVM, adaboost classifier, and bagged random forest, as well as two different variants of deep learning model RNN known as LSTM and Bi-directional LSTM [6]. Their model was specifically applied to analyze handwritings from patients with PD. In this paper, we will explore the performance of our novel pipeline model on vocal biomarkers, a different yet equally important domain for PD diagnosis.

### 1.4 Explainable AI

SHAP has been effectively implemented to explain various model outputs for diagnosing conditions like myocardial infarction (MI). Salih et al. applied SHAP to 4 classification models and generated plots similar to Figure 7a. According to SHAP, all 4 models agreed that high cholesterol, hypertension, and sex were the three most important factors determining an MI diagnosis, thus proving SHAP’s success [7]. We intend to use SHAP to identify important patterns in voice data that influence a PD diagnosis the most. By transparently quantifying the impact of each feature, our model will promote greater trust among clinicians and patients regarding the AI diagnostic process, setting this project apart from less interpretable models.

### 1.5 Current challenges

One major challenge is that while deep learning models have achieved high precision levels, most lack data explainability. This is particularly concerning in medical contexts where understanding the decision-making process of AI is crucial for gaining trust among healthcare professionals and patients [8]. Furthermore, the generalizability of these models across diverse populations is limited because they are only trained on specific demographic groups’ audio recordings. To enhance their robustness, there is a need for diverse datasets and training across various cohorts [9]. “Today, the much-needed personalization of medicine for PD patients still depends largely on the abilities, experience and intuition of treating physicians, nurses and allied healthcare professionals to adjust evidence-based medicine to individual decision making” [10]. This paper will utilize a large language model (LLM) to attempt to provide explainable AI that could personalize PD treatment.

### 1.6 Research aims

1. Automatic detection and prediction of PD binary classification using vocal biomarkers.
2. Use explainability to identify latent and important patterns in the input data in retrospect to the target to inform the definition of Parkinson’s through voice characteristics.
3. Generate a probability to create a scoring system of a patient’s odds of PD as a spectrum.

## 2 Methods

### 2.1 Data preprocessing

The dataset for training this AI model consists of 81 distinct voice recordings sourced from a publicly accessible dataset. Of these recordings, 41 were taken from healthy patients in the HC group, and the other 40 were taken from patients with PD who comprise the PD group. To maintain consistency among the data, the recordings were modified to remove background noise, equalize decibels based on sex, and retain intervals of silence before and after the audio.

### 2.2 Patient demographics

Iyer, A., et al. created the dataset shown in the Table 1a [11].

**Table 1a.**
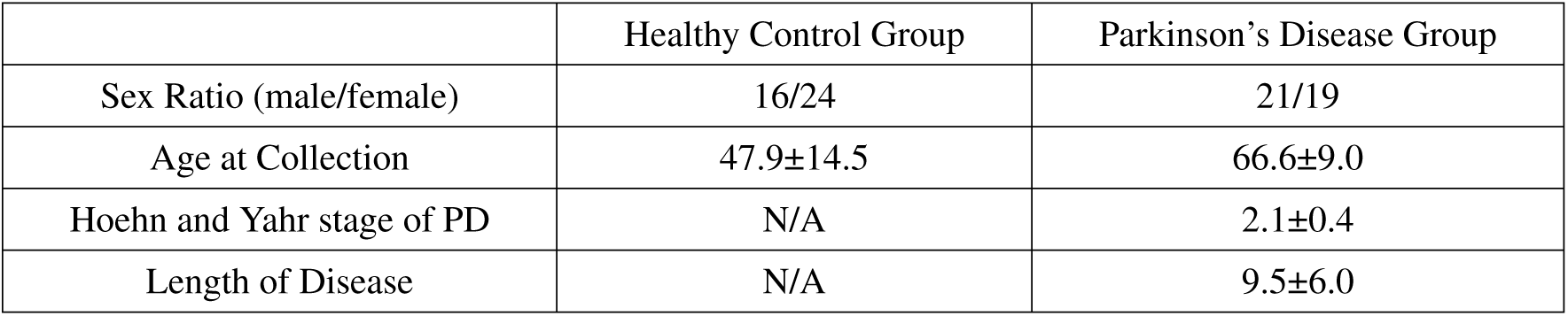

### 2.3 Data analysis

The AI model excels at processing audio files, demonstrating superior performance with `.wav` formats and `.zip` archives. The script uses the Parselmouth library, a Python wrapper for Praat—a software tool for speech analysis. The primary function, `extract_voice_features`, takes an audio file as input and extracts several key acoustic features. First, the audio is converted into a `parselmouth.Sound` object, allowing for various analyses. Then, using Praat’s “To Pitch” method, the AI retrieves the mean, minimum, and maximum pitch values. Next, the model calculates local jitter, which measures frequency variation, by converting the sound to a point process and applying the “Get Jitter (local)” method. Similarly, local shimmer, which measures amplitude variation, is extracted using the “Get Shimmer (local)” method. Finally, the script calculates the harmonicity-to-noise ratio (HNR) using Praat’s harmonicity analysis method to determine the mean HNR.

Figure 1a showcases an amalgamation of each acoustic feature extracted and collected.

**Figure 1a.**
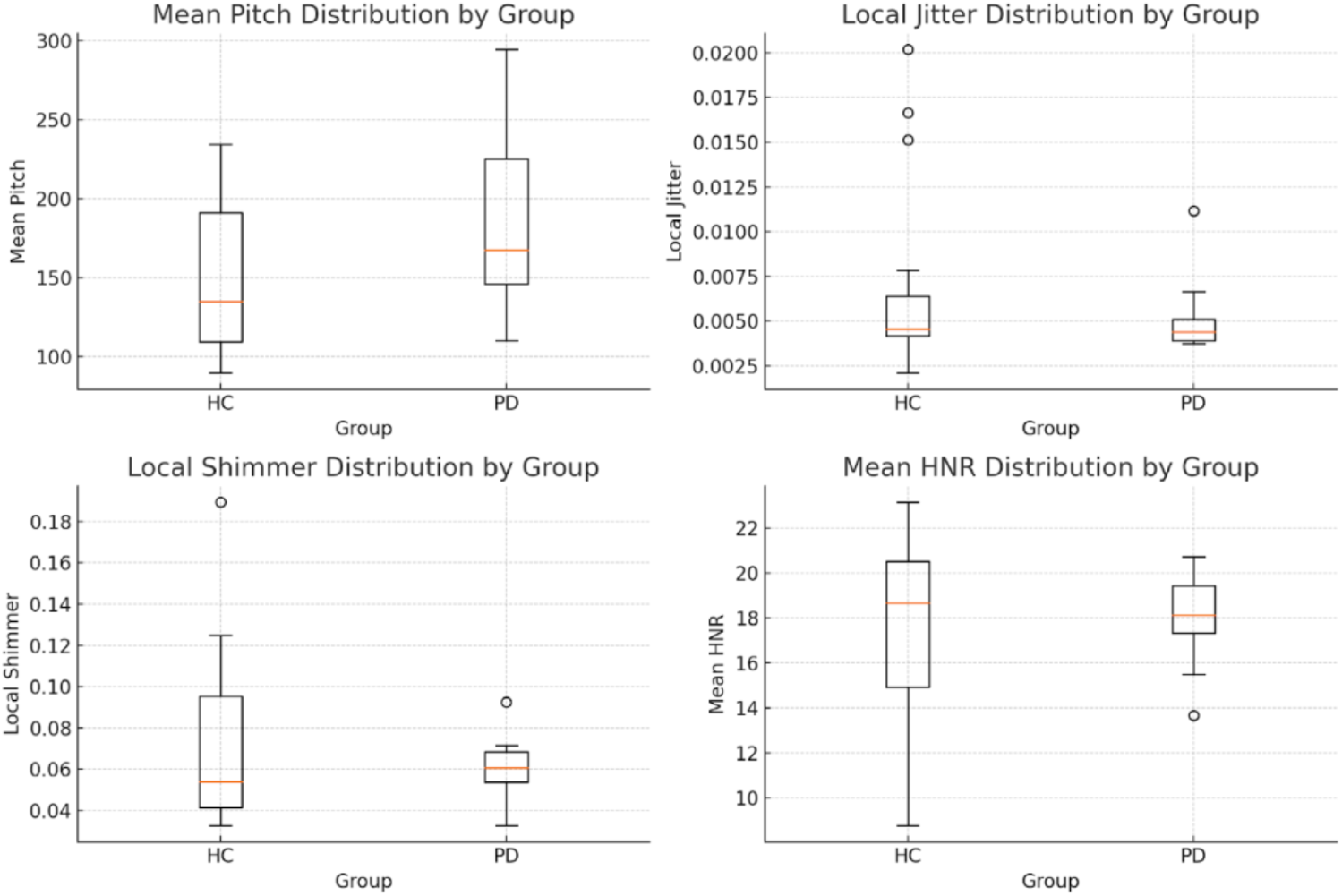

Raw data values can be found in the following document:

https://docs.google.com/document/d/1cW306roQFkzHqXWQPUEKz06Wbkcb89dGPkILhVwLrEA/edit?usp=sharing

Continuous model refinement involved leveraging insights from acoustic feature analysis and interpreting various graphical plots. Although these graphical visualizations were not directly used in the model, they served as a valuable tool for clinicians and researchers to better understand the underlying data. For example, the spectrograms provided visual cues about the voice recordings’ frequency content and temporal dynamics. Violin plots, box plots, and histograms illustrated the distribution of the acoustic features, highlighting blatant and nuanced differences between the HC and PD groups^1^. Scatter plots depicted relationships between mean pitch and HNR, revealing distinct clusters of the two groups, further aiding the model’s training. Overall, the graphical data allowed for a more informed approach to model improvement by enhancing our understanding of the data’s complexities.

### 2.4 Fourtier transformation

The Fourier transform (FT) is beneficial in speech analysis because different aspects of the voice can be analyzed more effectively in the frequency domain. Furthermore, the precise data of acoustic feature analysis is too complex for human scrutiny (Figure 2a vs. Figure 2b), especially in real-time. This gives a legitimate case for using machine learning to ensure proper analysis.

**Figure 2a.**
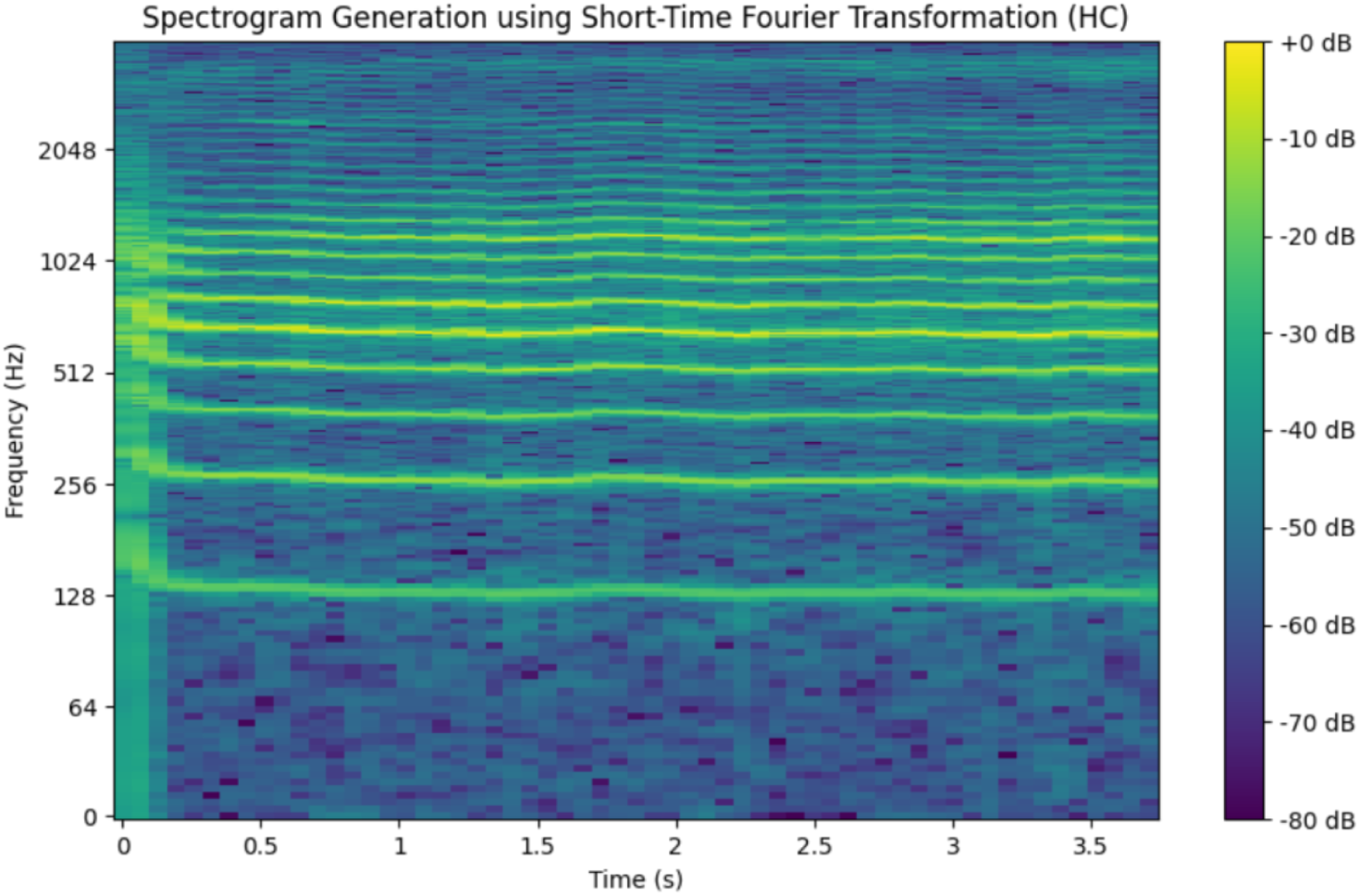

**Figure 2b.**
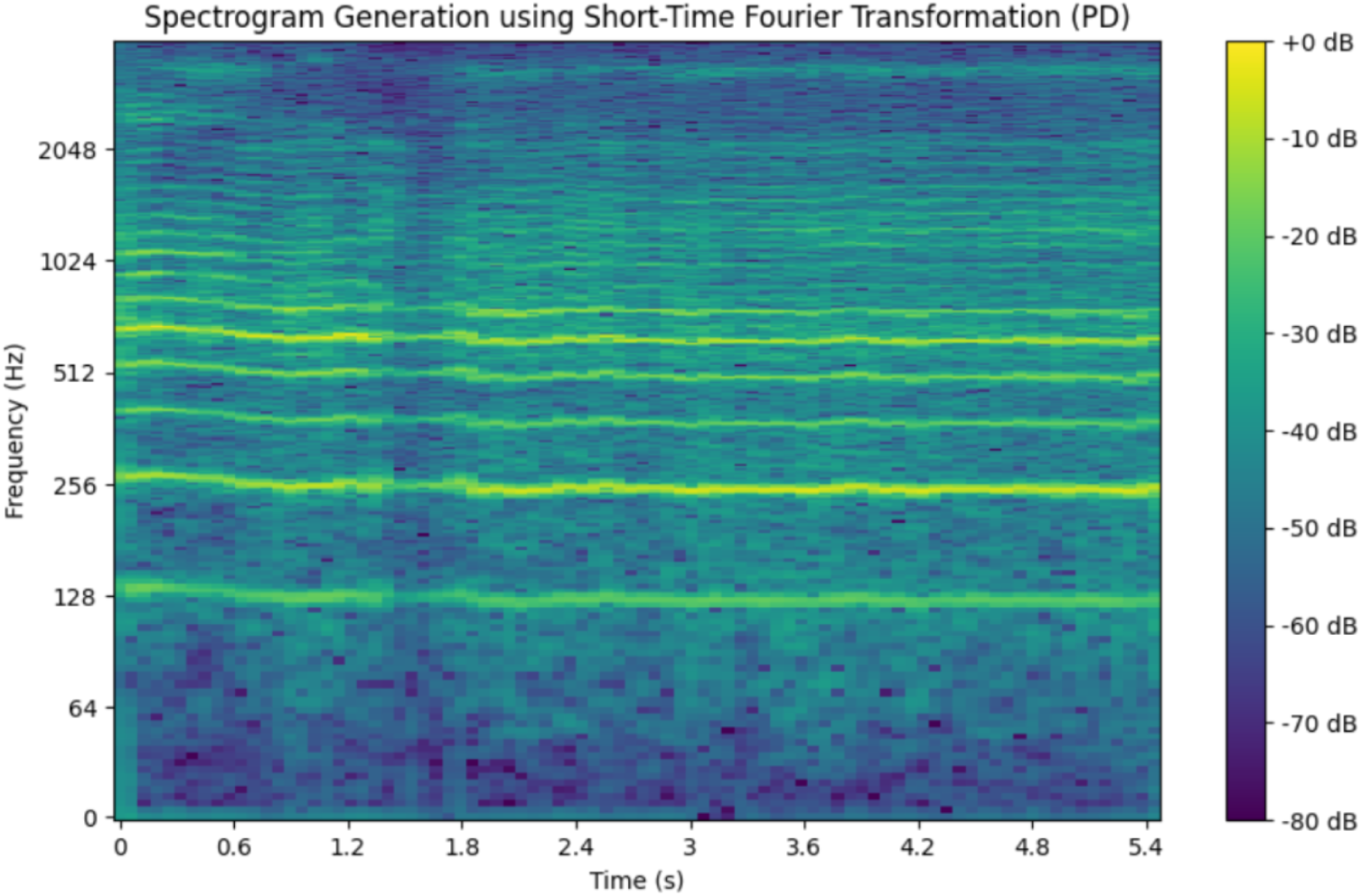

#### 2.4.1 Convertion to frequency domain

The 81 speech recordings were initially captured as time-domain signals, representing how the audio amplitude varies over time. The FT converts these time-domain signals into the frequency domain, representing the signal in terms of its frequencies and their respective amplitudes. This helps to isolate and identify different frequency components indicative of vocal characteristics.

#### 2.4.2 Feature extraction

The frequency domain representation obtained through FT allows the AI to extract the key acoustic features: pitch, jitter, shimmer, and HNR. The extracted features are then standardized using Python’s ‘StandardScaler’ from ‘sklearn’ to ensure they are on a similar scale, improving the model’s performance. The AI is trained to automate these extractions, allowing future researchers to efficiently process large datasets of voice recordings while maintaining consistency in feature extraction.

#### 2.4.3 Data visualization: spectrogram generation

The AI model generates spectrograms by applying the Short-Time Fourier Transform (STFT) to the audio signal. The STFT divides the signal into short, overlapping segments and then applies FT to each segment, resulting in a time-frequency representation. The colour scale adjacent to each spectrogram represents the magnitude of its frequency components, which are measured in decibels. The colours range from bright yellow, signifying higher amplitude components, to dark purple, indicating lower amplitude components. The colour scale represents 10 log(|S|/max(|S|)), where S denotes the complex numbers obtained from the output of the FT. This logarithmic scaling underscores the differences in intensity across various frequencies, allowing the AI model to easily discern subtle variations that could be crucial for diagnostic purposes.

#### 2.4.4 Spectrogram analysis: HC vs. PD

In Figure 2a, which depicts the HC group, the frequency bands are clearly defined and consistent across the time axis, indicating stable vocal tract function and regular vocal fold vibration. Furthermore, harmonics are visible at regular intervals—characteristics of a healthy vocal system. On the other hand, the PD group in Figure 2b has less distinct frequency bands and exhibits more variability. These irregularities suggest vocal instability, something commonly seen in PD patients. Such instability may be due to tremors affecting vocal fold vibration, leading to the scattered and less defined harmonic structure. We only used vocal data in our AI decision-making, but these graphs exist as a foundational representation to aid clinicians in understanding the granular details of the voice recordings.

### 2.5 Experimentation

The experimentation phase of this model involved constantly improving a rudimentary MLP and CNN model designed to diagnose Parkinson’s disease from voice recordings. This model utilized Python’s robust ML and audio processing libraries for rigorous training and validation to achieve optimal performance. Eventually, MLP and CNN were paired with RNN and MKL to create a unified PD diagnosis model that harnesses each approach’s strengths [6].

#### 2.5.1 Model architecture and Training

The MLP + CNN + RNN + MKL (our champion model) is a hybrid model whose architecture was designed to discern intricate patterns in voice signals. MLP is useful when applied to structure learning, meaning it is strong when detecting and learning patterns in HC and PD recordings. CNN excels at capturing local acoustic patterns with spectrograms, such as mean pitch. RNN effectively models the temporal dynamics of speech, which is critical for the model to correctly identify sequential anomalies associated with PD. Finally, MKL enriches the AI model by enabling the integration of diverse feature modalities, thus making for a more comprehensive analysis and prediction. This combination not only advanced model precision, sensitivity, and recall but also enhanced generalizability across different data sets. The model comprises multiple convolutional layers that extract hierarchical feature representations from the input spectrograms. These layers were followed by pooling layers to downsample the feature maps, reducing computational complexity and preventing data overfitting. Finally, dense layers were used to combine the extracted features and make a final prediction. We used k-fold cross-validation (CV) to report more accurate evaluation results, which means we averaged final performance metrics across all runs.

This model can be considered a sequential pipeline structure with multiple branches where CNN and RNN might operate in parallel, and their outputs are later combined using MKL [6].

#### 2.5.2 Architecture flow

1. **Input Data:**

**•** Input is fed into the CNN layers.
2. **CNN Layer**

**•** Extracts feature maps from the input data.
**• Output:** Feature maps.
3. **RNN Layer**

**•** Takes feature maps as input and learns temporal sequences.
**• Output:** Temporal feature representation.
4. **MKL Layer**

**•** Takes inputs from both CNN and RNN.
**•** Learns a kernel-based representation.
**• Output:** Combined representation.
5. **MLP Layer**

**•** Processes the combined representation to learn higher-level, non-linear relationships between acoustic features.
**• Output:** A refined, non-linear representation of the data suitable for prediction.
6. **Fully Connected Layers**

**•** The final representation is passed to one or more dense layers.
**• Output:** Prediction distribution.
7. **Output Layer**

**•** Provides the classification output.

**Figure 3a.**
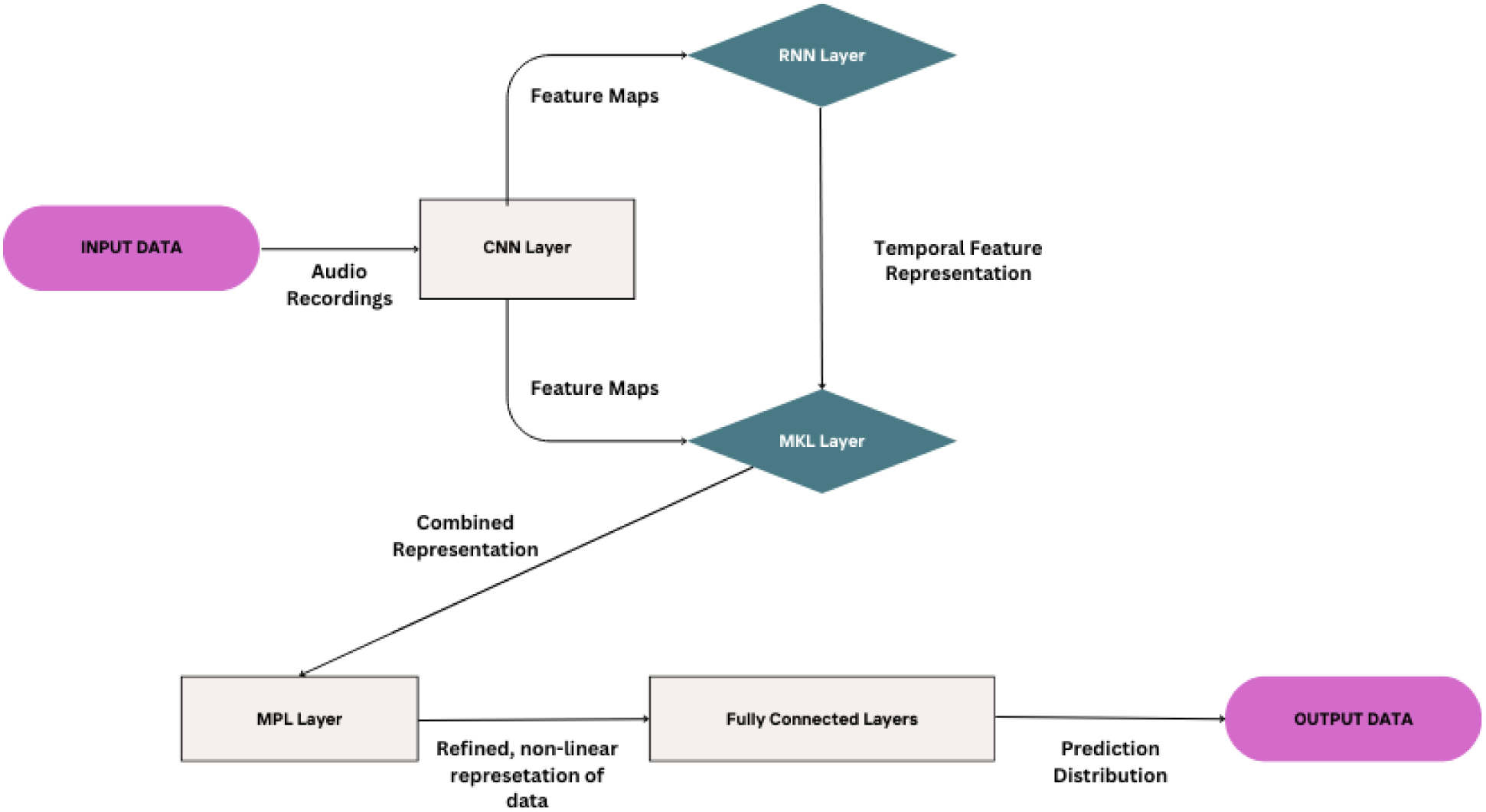

### 2.6 Scoring system

#### 2.6.1 Introduction of the scoring system

HC and PD diagnoses exist on a continuum, meaning that not all files labeled as “HC” are equally distant from a potential PD diagnosis; some may be closer to being classified as PD than others. In conjunction with binary labeling, we want a more granular means of distinction among different HC and PD cases.

We created a scoring system to quantify the likelihood of a patient having PD based on key acoustic features extracted from their voice recordings. This system is derived from model probabilities, allowing clinicians to interpret the likelihood of PD more effectively and set individualized thresholds for diagnosis. By enabling precision medicine, this approach ensures that diagnostic decisions are tailored to the unique characteristics of each patient rather than relying on a one-size-fits-all binary system.

#### 2.6.2 Description of the scoring system

Our scoring system assigns probabilities of the likelihood of an individual being diagnosed with PD on a scale from 0-1. The system uses probabilities generated by a Random Forest model trained on acoustic vocal features such as mean pitch, MFCCs, local jitter, local shimmer, and HNR. This model is integrated in a sequential pipeline with our champion model. This means that once the champion model completes its diagnosis, the results are input into the Random Forest model for scoring. Although the Random Forest model’s code is computationally simpler than our champion model’s code, it shows a strong correlation with the champion model’s results, thus providing an interpretable layer for clinical decision-making ^2^.

## 3 Results

This section will report the model evaluation results. The primary metrics for model evaluation were accuracy and cross-entropy loss, which were assessed during both the training and validation phases. Accuracy indicates how well the model correctly predicts the inputted data’s labels. A high accuracy indicates that the model can adequately distinguish between HC and PD recordings. On the other hand, low accuracy suggests a higher number of misclassifications. Cross-entropy loss measures how well or poorly the model’s predictions match the actual labels during training. A high cross-entropy loss value (40% or higher) indicates the predictions significantly deviate from the actual labels. In contrast, a cross-entropy low loss value (20% or lower) shows the predictions are closely aligned with the actual labels. We utilized a 5-fold CV in which the stratified data was split into 5 subsets. Each fold further trains the model on 4 subsets and validates it on the remaining subset. This process was repeated for each of the 5 folds, ensuring that every data point was used for training and validation to report a consistent average of evaluation indices. We also ensured that each subset of the data was used for validation precisely once. This approach mitigates the risk of overfitting and provides a more reliable estimate of the model’s performance.

The most optimal model was the MLP + CNN + RNN + MKL model. Its performance was evaluated based on accuracy, precision, recall, F1 score, and AUC metrics. Its average accuracy was 0.9111, indicating that around 91.11% ± 1 of all predictions made with this model will be correct. For reference, a 2016 meta-analysis of 11 pathologic examinations (the gold standard for PD diagnosis) for PD had a pooled diagnostic accuracy of 80.6% [12]. The precision was 0.8984, meaning roughly 89.84% ± 1 of the positive predictions (Parkinson’s disease) were correct. This high precision value indicates that the model is reliable when it predicts a patient with Parkinson’s, as it produces a low rate of false positives. This low rate of false positives can save patients and hospitals resources by preventing healthy patients from testing positive for Parkinson’s. It will also prevent false anxiety from being instilled within the patient. The recall was 0.9250, indicating that the model’s ability to identify positive cases correctly was 92.50% ± 0.5. The F1 score, which was 91.13% ± 0.1, balances high precision and recall. This score reflects the model’s ability to accurately identify Parkinson’s patients while keeping false positives to a minimum. The MLP + CNN + RNN + MKL model outperforms all other models on every metric.

The loss values of our champion model remained consistently low, as seen in Figure 4b. The loss value ranged from a high of 9.88% in Folds 4 and 5 to a low of 7.41% in Fold 3. The average loss value of the champion model is 8.89% ± 1.

Our champion model’s consistency in both accuracy and cross-entropy loss shows that it is extremely good at predicting unseen data.

**Figure 4a.**
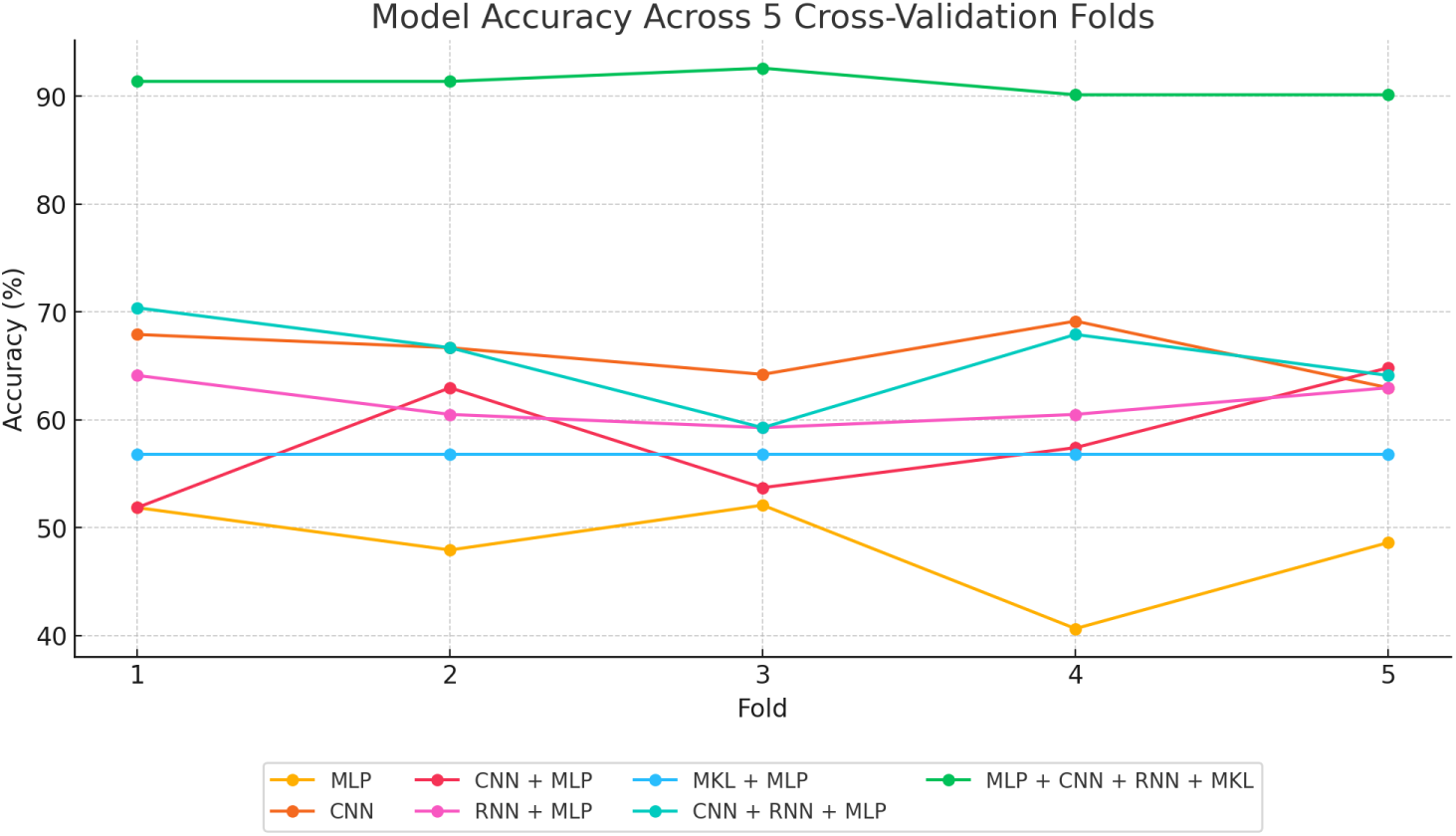

**Figure 4b.**
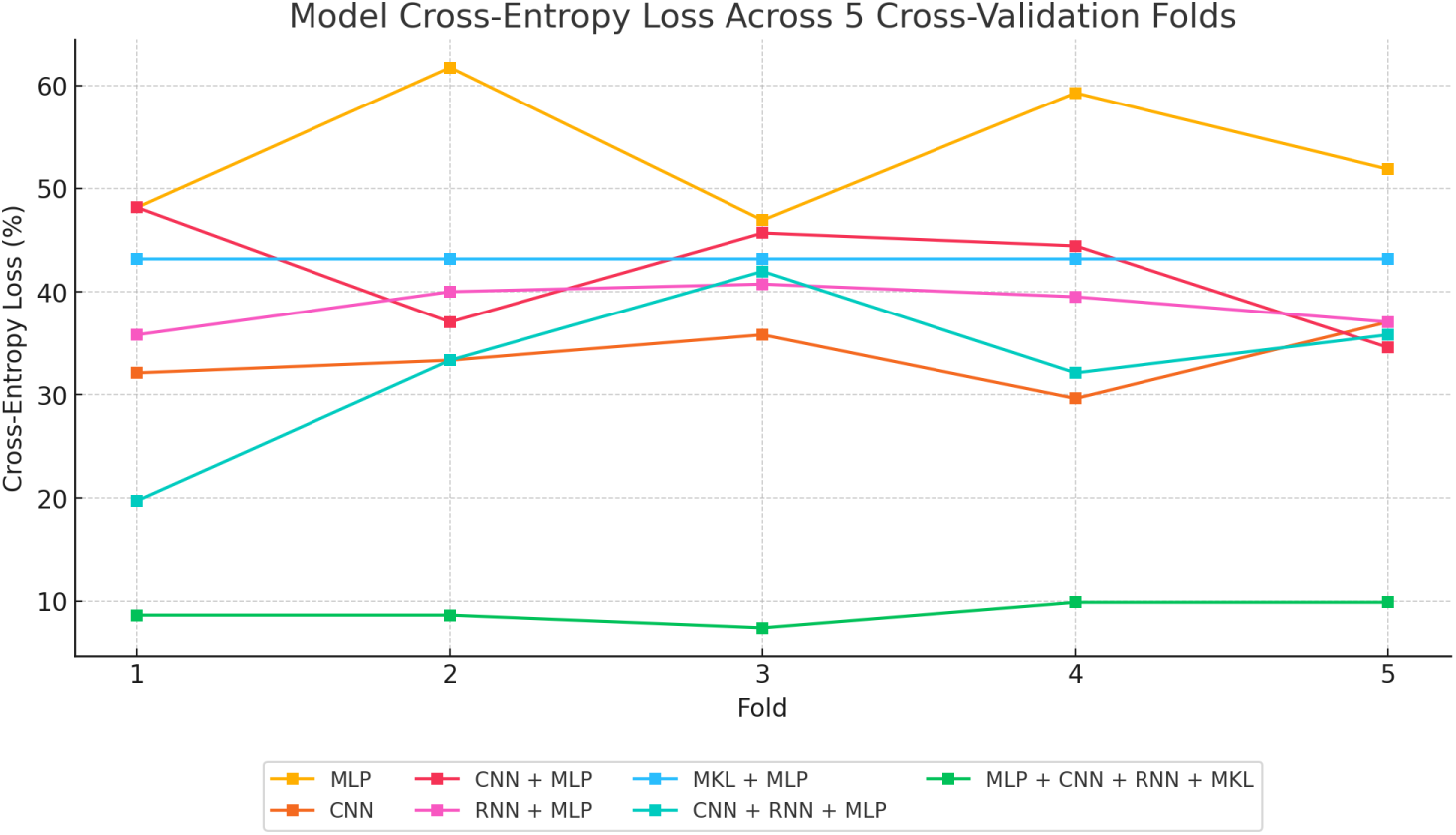

**Figure 5a.**
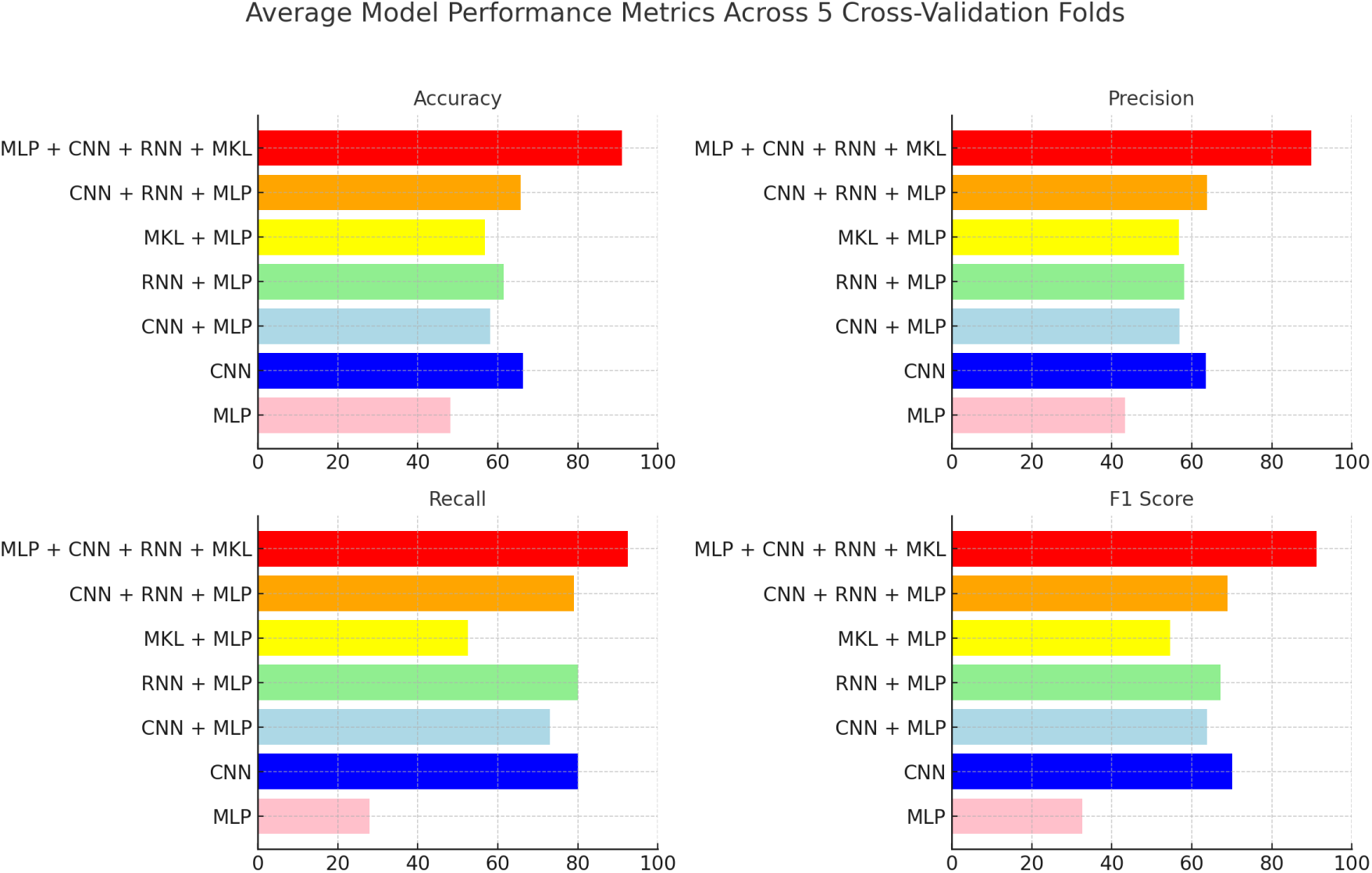

The Receiver Operating Characteristic (ROC) shows the trade-off between the true positive rate (TPR) and false positive rate (FPR) for different machine learning models. The higher the AUC, the better the model is at distinguishing between positive and negative cases. The dashed line represents random chance (AUC = 0.5), and models performing above this line indicate better-than-random performance.

**Figure 6a.**
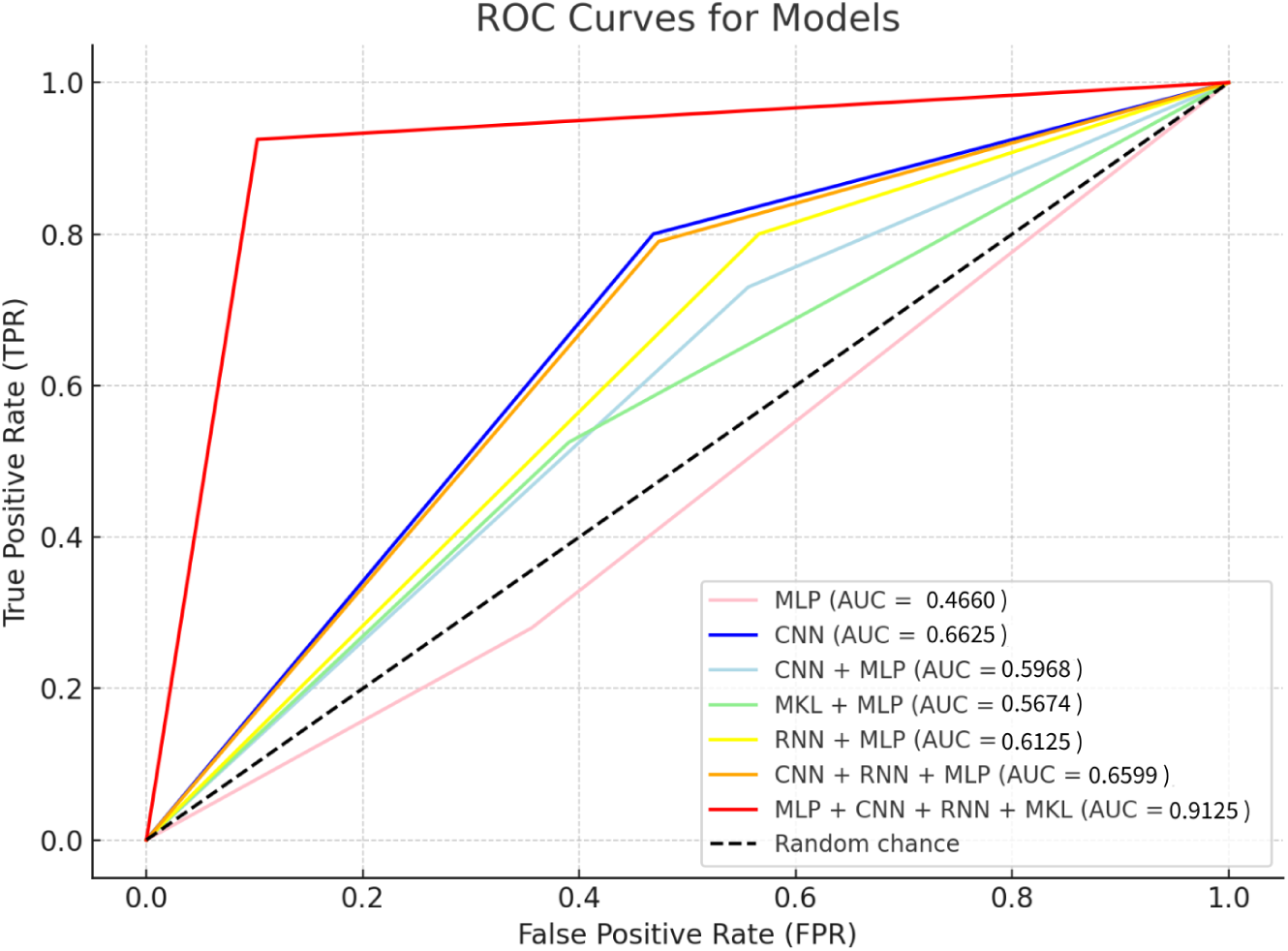

Our champion model achieved an AUC value of 0.9125, as shown in Figure 6a. This means it has strong discriminative power when distinguishing between individuals with Parkinson’s disease and healthy individuals across different classification thresholds. This value is comparable to Iyer et al. [11]. They used a CNN with transfer learning approach on the same 81 audio files we used. However, they did not report accuracy, recall, precision or F1 score.

### 3.1 Scoring system results

#### 3.1.1 Applying the scoring system to our data

The outcomes of the implemented scoring system demonstrate a distinct separation in the probability assessments for PD across the 81 analyzed audio samples. There is a clear demarcation of which files were considered HC and PD based on the system. For example, 40 of the 41 HC files scored between 0-0.30. However, File `AH_678A_2E7AFA48-34C1-4DAD-A73C-95F7ABF6B138.wav`, classified as HC, was assigned a higher score of 0.39. According to Table 2a, this file has a moderate likelihood of developing PD, suggesting that such a case would require careful monitoring in a clinical setting. Conversely, 38 of the 40 PD files scored between 0.70-0.90. Notably, the files `AH_545812846-0C14B32A-6C50-4B62-BC89-0A815C2DEEFA.wav` and `AH_545880204-EE87D3E2-0D4C-4EAA-ACD7-C3F177AFF62F.wav` registered scores of 0.69 and 0.62, respectively. Upon further analysis of the files scoring 0.39 and 0.62, their acoustic features closely resemble those of patients in the early stages of PD. This observation validates our scoring system by confirming that the vocal biomarkers in the audio files accurately correspond with their assigned scores.

**Table 2a.**

| Scoring Range | Description |
| --- | --- |
| 0.00 - 0.10 | Very low likelihood of PD. Healthy vocal features with no signs of PD. |
| 0.10 - 0.20 | Low likelihood of PD. Some vocal features may be slightly atypical but are generally not indicative of PD. |
| 0.20 - 0.30 | Mild likelihood of PD. Features start to show more noticeable deviations, suggesting further monitoring and evaluation. |
| 0.30 - 0.40 | Moderate likelihood of PD. This range suggests that several vocal features are indicative of early signs of PD and should be closely monitored. |
| 0.40 - 0.50 | Moderate to high likelihood of PD. There is a significant indication of Parkinson's based on vocal features, suggesting a need for clinical evaluation. |
| 0.50 - 0.60 | High likelihood of PD. Vocal features strongly indicate PD, and clinical assessment is strongly recommended. |
| 0.60 - 0.70 | Very high likelihood of PD. Most vocal features are consistent with those observed in PD patients. |
| 0.70 - 0.80 | Extremely high likelihood of PD. Vocal features are highly indicative of advanced characteristics of PD. |
| 0.80 - 0.90 | Near certainty of PD. Almost all vocal features align closely with known PD patterns. |
| 0.90 - 1.00 | Definite likelihood of PD. The vocal features meet all or nearly all the criteria for PD according to data from our AI model and existing literature. |

On the more definitive end, File `AH_322A_C3BF5535-A11E-498E-94EB-BE7E74099FFB.wav` was scored at 0.06, indicating a virtually nonexistent likelihood of PD, and File `AH_545789670-C297FD53-BF71-4183-86A0-58E5E1EB0DF8.wav` received a score of 0.89, strongly suggesting PD presence. Subsequent analyses confirmed that their acoustic features are highly representative of their respective scores, thereby validating our scoring system even in extreme cases.

All results can be found in Table 4a by clicking the link:

**Table 3a.** 

https://docs.google.com/document/d/1-FS9LavZZTZEEBWXr3PPWISeQPDPUXM1j-vdNQA79dY

## 4 Discussion

In this juncture, we want to where the machine misclassified the predictions. We would also want to generate insights if possible to inform medical practitioners in the diagnosis and prognosis of PD using voice. We will also use an LLM to investigate the important vocal characteristics in the data.

### 4.1 Error analysis (champion model)

#### 4.1.1 Healthy control group misclassifications

Of the 41 HC audio files, an average of 36.8 were correctly classified as HC, while 4.2 were incorrectly classified as PD. This resulted in a precision rate of 89.84% ± 1 for HC label predictions. The misclassification of an average of 4.2 HC files could be attributed to various reasons. The most likely explanation for the observed overlap in acoustic features between HC and early-stage PD patients can be attributed to the subtler distinctions between these groups compared to those between HC and late-stage PD. This nuance results in the model occasionally misclassifying HC files as PD due to their acoustic similarity. However, it is also possible the feature extraction software did not adequately capture variations in speech patterns, thus resulting in too much leniency in the model’s decision boundary when distinguishing between the two classes.

#### 4.1.2 Parkinson’s disease group misclassifications

The AI model correctly classified 37 out of the 40 PD files, with 3 files incorrectly classified as HC. This makes the recall rate 92.50% ± 0.5 for PD predictions, indicating that few PD instances were missed. These false negatives are particularly concerning in a clinical context because failure to identify PD could delay treatment. The variability in symptom severity among patients may have contributed to the misclassification of PD. The model may have also been overly conservative when labeling borderline cases as PD, resulting in such prediction errors.

Table 5a represents the average actual frequency values of the seven AI Models tested across 5 CV folds. Each model was evaluated on the original 41 HC files and the original 40 PD files. A higher count in the “Correctly Predicted” section indicates stronger model performance.

**Table 5a.** represents the average actual frequency values of the seven AI Models tested across 5 CV folds. Each model was evaluated on the original 41 HC files and the original 40 PD files. A higher count in the “Correctly Predicted” section indicates stronger model performance.

| Model | HC |  | Total HC Files | PD |  | Total PD Files | Total Correct | Total Incorrect |
| --- | --- | --- | --- | --- | --- | --- | --- | --- |
|  | Correctly Predicted | Mistaken For PD |  | Correctly Predicted | Mistaken For HC |  |  |  |
| MLP | 26.4 | 14.6 | 41 | 11.2 | 28.8 | 40 | 37.6 | 43.4 |
| CNN | 21.8 | 19.2 | 41 | 32 | 8 | 40 | 53.8 | 27.2 |
| CNN + MLP | 18.2 | 22.8 | 41 | 29.2 | 10.8 | 40 | 47.4 | 33.6 |
| MKL + MLP | 25 | 16 | 41 | 21 | 19 | 40 | 46 | 35 |
| RNN + MLP | 17.8 | 23.2 | 41 | 32 | 8 | 40 | 49.8 | 31.2 |
| CNN + RNN + MLP | 21.6 | 19.4 | 41 | 31.6 | 8.4 | 40 | 53.2 | 27.8 |
| MLP + CNN + RNN + MKL | 36.8 | 4.2 | 41 | 37 | 3 | 40 | 73.8 | 7.2 |

### 4.2 Feature importance explainability

In this study, we employed SHAP to interpret the model’s predictions. SHAP generations offer insight into the extent to which each feature contributed to the final predictions, allowing us to validate the model’s decision-making process and reliability.

#### 4.2.1 Key features and their contributions

The SHAP summary plot (Figure 7a) provides a thorough visualization of the most influential features used by our pipelined composite champion model to distinguish between HC and PD patients. Each feature’s impact on the data output is displayed along the x-axis. Positive SHAP values indicate a higher likelihood of the prediction being PD, and negative values indicate a higher likelihood of HC. The left-hand-side y-axis shows the features that had the most influence on model output (top) and the least influence on model output (bottom).

#### 4.2.2 Mel-frequency cepstral coefficients (MFCC) features

Among the most impactful features were MFCCs, with `mfcc_3`, `mfcc_11`, and `mfcc_5` showing significant influence (Figure 7a). MFCCs encapsulate the spectral properties of voice, which are known to be altered in PD patients due to the neurodegenerative nature of the disease on speech production. The efficacy of MFCCs in speech recognition, speaker biometry, or voice pathology detection is universally recognized [13]. In fact, a 2023 study isolated MFCCs from other speech features and analyzed sustained vowels similar to this paper. Their performance metrics ranged from 70 to 79%, highlighting the relevance of MFCCs in this field [14].

Notably, `mfcc_3` had a strong positive SHAP value, signifying that higher values of this feature were associated with an increased likelihood of a PD diagnosis. An absence of `mfcc_3` would indicate a likelihood of HC. Some characteristics residing at the center of Figure 7a, such as `mfcc_2`, are more neutral when predicting both values of our binary target. Other characteristics such as `mfcc_6`, which is found at the bottom of Figure 7a, indicate less prominence in model decision-making with reference to either values of our binary target.

#### 4.2.3 Jitter and shimmer

Incorporating jitter and shimmer measurements provided deeper insight into the fine vocal variations associated with PD. Local shimmer and local jitter were extremely influential because they allowed the model to recognize sensitivity in amplitude and frequency variations. For instance, high values of local shimmer were linked to a higher likelihood of a PD diagnosis, as shown by the positive SHAP values. Similarly, `rap_jitter` and `local_jitter`, which measure relative frequency perturbations, were also crucial in the model’s predictions.

#### 4.2.4 Harmonicity-to-noise ratio

Although not as significant, ‘mean HNR’ still contributed to the models’ predictions. Lower HNR values, which suggest a noisier voice signal, were more often associated with PD. This supports the clinical observations of PD patients tending to have a breathier voice due to impaired control of vocal fold vibration [15].

**Figure 7a.**
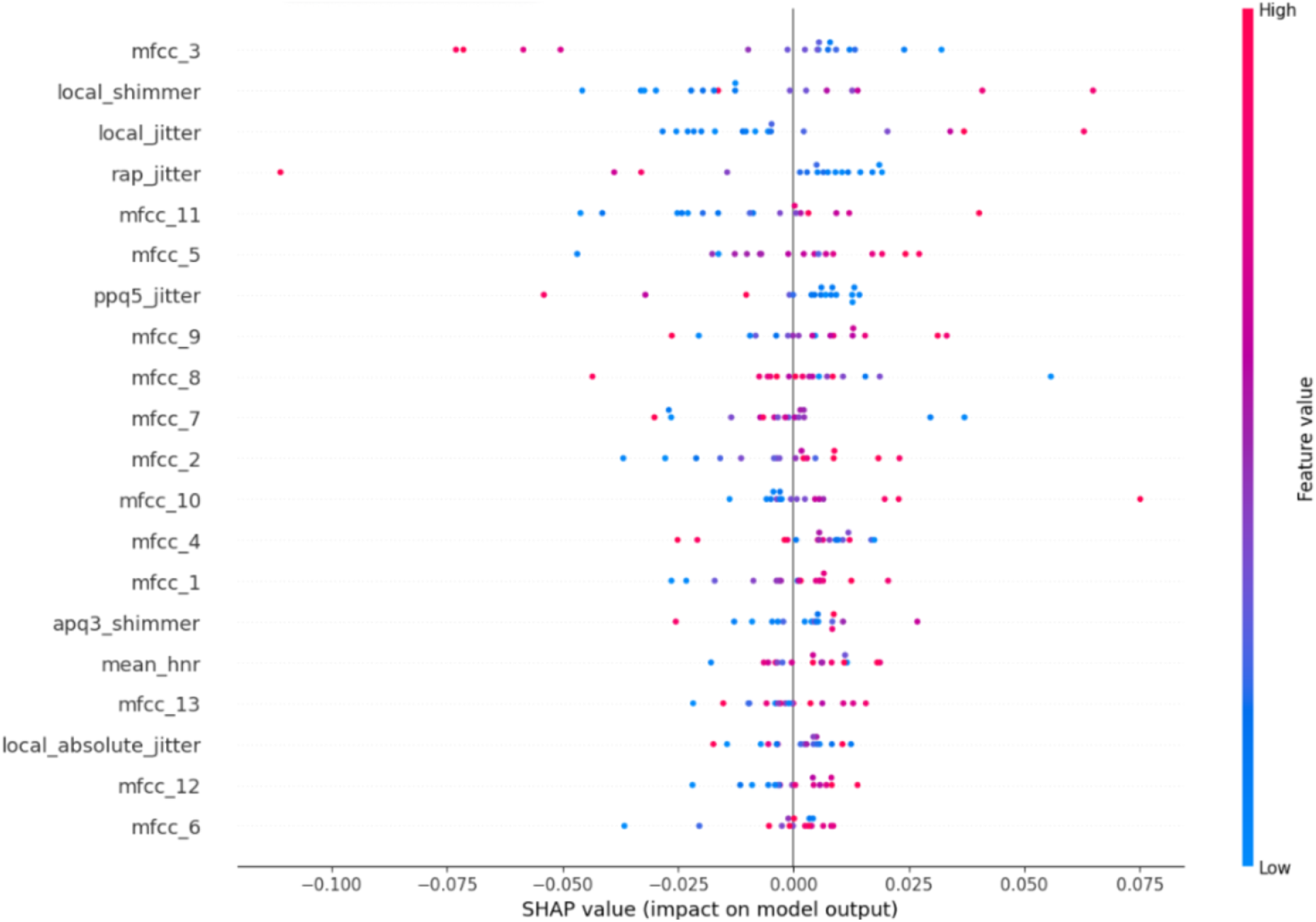

## 5 Limitation

Due to the small sample size of 81 audio files, the results may not reflect the model’s true capabilities and undermine generalizability. In the continuation of this project, we will continue collecting data to improve the model’s training, thus improving generalizability. The scarcity of available data also potentially magnifies the biases of age and gender depicted in Table 1a. Furthermore, the PD data includes a mix of early and late-stage PD files, thus skewing the model’s ability to discern between files.

## 6 Suggestions for future research

Using MLP + CNN + RNN + MKL in the AI model introduced several layers of complexity. While highly sophisticated and extremely powerful, this model style may have introduced new layers of depth that were not properly synthesized, thus potentially contributing to data overfitting or poor generalization for specific test cases. This warrants future work to develop methods of balancing the complexity of the AI with the data. The most logical way would be to train this AI with many more audio files and run breakages in epochs to prevent overfitting. Furthermore, it would be prudent to attempt to pair this MLP + CNN + RNN + MKL model with other means of physical analysis. For example, creating a smartwatch that can record the wearer’s speech and track physical movements such as tremors and gait would be a great way to introduce a multimodal, non-invasive, early PD diagnosis method.

## 7 Conclusion

This study highlights the efficacy of AI, particularly a hybrid model combining MLP, CNN, RNN, and MKL in diagnosing early PD through voice analysis. The model demonstrated a robust ability to distinguish between HC and PD patients with significant accuracy by leveraging key vocal biomarkers such as MFCCs, jitter, and shimmer.

Our champion model had an accuracy of 91.11%, a precision of 89.84%, a recall of 92.50%, an F1 score of 91.13%, and an AUC of 0.9125. These evaluation metrics are all around the 90% mark, indicating high consistency in distinguishing PD patients.

Furthermore, the use of SHAP for data explainability reinforced the reliability of the diagnostic tool by providing transparent insight into how individual acoustic features impacted model decision-making. Features like MFCCs have been well-documented in existing literature as strong indicators of vocal abnormalities in PD, which is why they were among the most prominent in Figure 7a. Also, jitter and shimmer significantly contributed to model decision-making, aligning with well-tested clinical characteristics of PD-related speech disorders. Extrapolating from just the raw data, LLMs such as SHAP can provide insights that were otherwise latent, potentially enabling physicians to tailor treatment plans more effectively by identifying the most prominent acoustic features in a patient’s voice data. In Figure 7a, for instance, features such as `mfcc_3` or `local_shimmer` are more pronounced, indicating different aspects of disease progression that can guide individualized treatment planning.

Also, implementing a scoring system proves advantageous over similar works because it allows for a quantifiable, objective measurement of disease markers, which is crucial for early diagnosis and management of PD. Using a random selection of voice recordings, we validated our scoring system and it was consistent with the prediction results because the HC voice recordings were scored 0-0.40, and the PD voice recordings were scored 0.60-0.90, which are the correct ranges for the HC and PD recordings. This system facilitates longitudinal monitoring of disease progression, offering a valuable tool for assessing treatment efficacy and adjusting therapeutic interventions accordingly.

This study’s findings suggest that ML, coupled with a hybrid variation of advanced voice feature analysis, offers a promising and noninvasive approach for early PD diagnosis.

## 9 Conflict of interets

The authors have no conflict of interest to report.

## 10 Data availability

The data supporting the findings of this study are openly available in Figshare at https://doi.org/10.6084/ `m9.figshare.23849127`^3^. These data were derived from the following resources available in the public domain: https://figshare.com/articles/dataset/Voice_Samples_for_Patients_with_Parkinson_s_Disease_and_Healthy_Controls/23849127.

## 11 Appendix

### A. Technical Details and Definitions

1. **Mel-Frequency Cepstral Coefficients (MFCCs):**

**• Definition:** MFCCs are coefficients that collectively represent the short-term power spectrum of a sound.
**• Application:** In this paper, MFCCs such as `mfcc_3`, `mfcc_11`, and `mfcc_5` were used to distinguish between HC and PD, with higher MFCC values often indicating PD.
2. **Jitter:**

**• Definition:** A measure of frequency variation from cycle to cycle in voice signals, indicating potential vocal fold instability.
**• Application:** Used to identify fine variations in vocal recordings that are symptomatic of PD.
3. **Shimmer:**

**• Definition:** A measure of amplitude variation from cycle to cycle used to detect issues in vocal fold function.
**• Application:** Used to identify fine variations in vocal recordings that are symptomatic of PD.
4. **Harmonic-to-Noise Ratio (HNR):**

**• Definition:** HNR is the ratio between harmonic components of the data and noise components. Lower HNR values indicate a breathier or noisier voice, which can be indicative of PD.
**• Application:** HNR contributed to the AI model’s predictions by looking for conventional symptoms of PD.
5. **Fourier Transformation (FT):**

**• Definition:** A mathematical technique that transforms a time-domain signal into its constituent frequencies, providing a frequency-domain representation of the signal.
**• Application:** This study used FT to convert voice recordings into the frequency domain, aiding in accurately extracting key acoustic features.
6. **SHapley Additive exPlanations (SHAP):**

**• Definition:** Provides a way to explain the output of machine learning by showing the contribution of each acoustic feature to the predictions.
**• Application:** This study used SHAP to interpret the model’s predictions, offering insight into the extent to which acoustic features contributed to the final diagnosis.

### B. Model Architecture and Training Process

1. **MLP + CNN + RNN + MKL Learning Model Architecture:**

**•** This model combines MLP for non-linear data representation, CNN for local pattern recognition, RNN for temporal sequence analysis, and MKL for integrating multiple feature modalities.
**•** Architecture Overview:

**– CNN Layers:** Extracted hierarchical feature representations from input spectrograms.
**– RNN Layers:** Modeled the temporal dynamics of speech to identify sequential anomalies.
**– MKL Layers:** Integrated diverse feature modalities, enhancing generalizability and robustness.
**– MLP Layers:** Processes the combined representation to learn higher-level, non-linear relationships between acoustic features.
**• Training Process:** The model was trained using k-fold cross-validation to avoid overfitting and ensure quality output. The training involved multiple epochs, continuous monitoring, and tuning based on loss and accuracy metrics.
2. **Parameters:**

**• Learning Rate:** Adjusted based on training output to optimize model convergence.
**• Epochs:** An epoch is one complete pass through the entire training dataset. Training a model for multiple epochs improves performance by allowing it to learn patterns. This study trained AI models on a scale of 1-150 epochs with the flexibility to halt training at any point to prevent overfitting.
**• Batch Size:** All AI models were trained on the same 81 voice recordings to ensure consistency in input, thus minimizing confounding variables.

### C. Data Preprocessing Steps

1. **Noise Reduction and Decibel Equalization:**

**•** Iyer, A. et al. (2023) removed all background noise from the 81 audio recordings. Furthermore, audio was equalized based on sex to maintain consistency across data [11].
2. **Handling Silent Intervals:**
  **•** Iyer, A. et al. (2023) retained intervals of silence before and after the 81 audio recordings to preserve the natural speech patterns of the participants [11].
3. **Feature Extraction:**
  **•** All audio files were processed using a Parselmouth library, a Python wrapper for Praat. Key acoustic features were extracted.

### D. Supplementary Data and Visualizations

1. **Spectrograms:**

`•` HC and PD Spectrograms: https://drive.google.com/drive/folders/ `1gVih3iqpJXzdn0O5B4YT4W6HkmX6rPB2?usp=sharing`
Short-Time Fourier Transformation (STFT) Enhanced Spectrograms: Figures 2a and 2b are STFT-enhanced spectrograms, showing improvement in clarity and diagnostic utility.
2. **Evaluation:**

**•** Accuracy and Cross-Entropy Loss Metrics: This metric provides the accuracy and loss for each fold in the 5-fold CV, highlighting variability and areas for further model improvement.
  **–** An accuracy value above 80% indicates high-quality performance regarding the ratio of the number of correct predictions (both true positive and true negative) to the total number of predictions (true positive, true negative, false positive, and false negative).
  **–** A **loss** value below 20% indicates high-quality performance regarding predictions matching with true labels.
**•** The following is how the performance metrics were calculated:

**– Accuracy:** 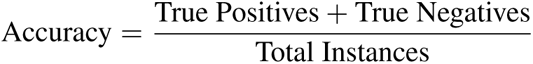
**– Precision:** 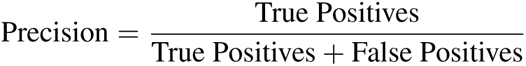
**– Recall:** 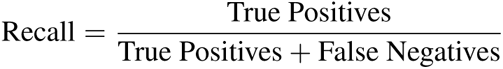
**– F1 Score:** 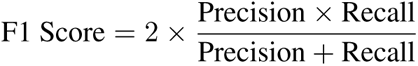
**– Confusion Matrix:** 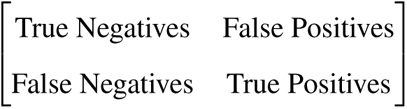
**–** The following table displays the average performance metrics across 5-fold CV of each model:

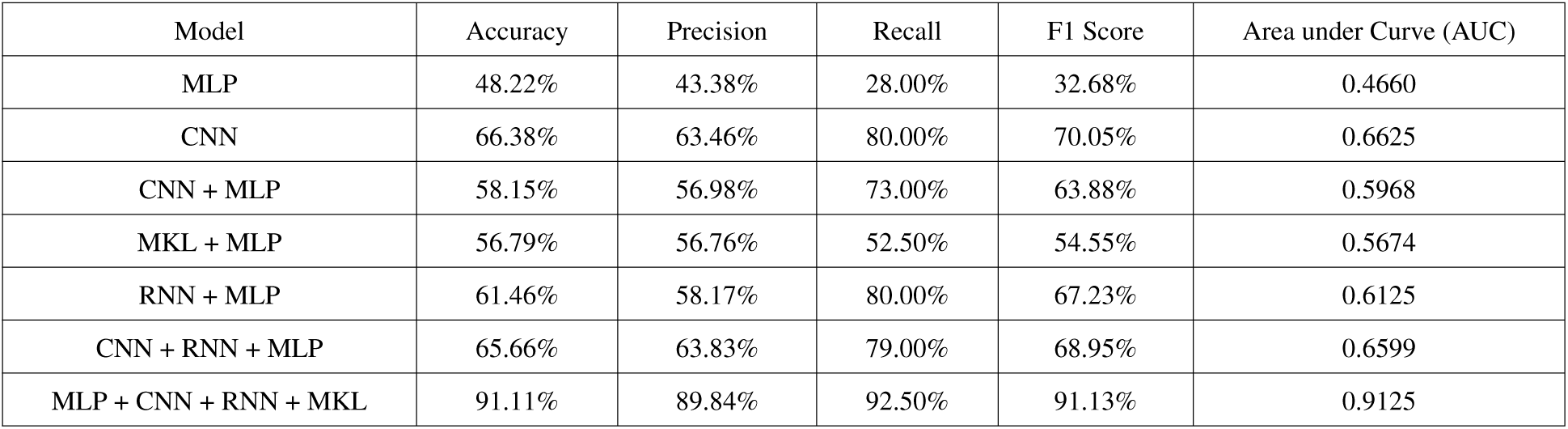

### E. Code and Algorithm Explanations

1. **Pseudocode for Model Training:**

**•** Model Workflow:

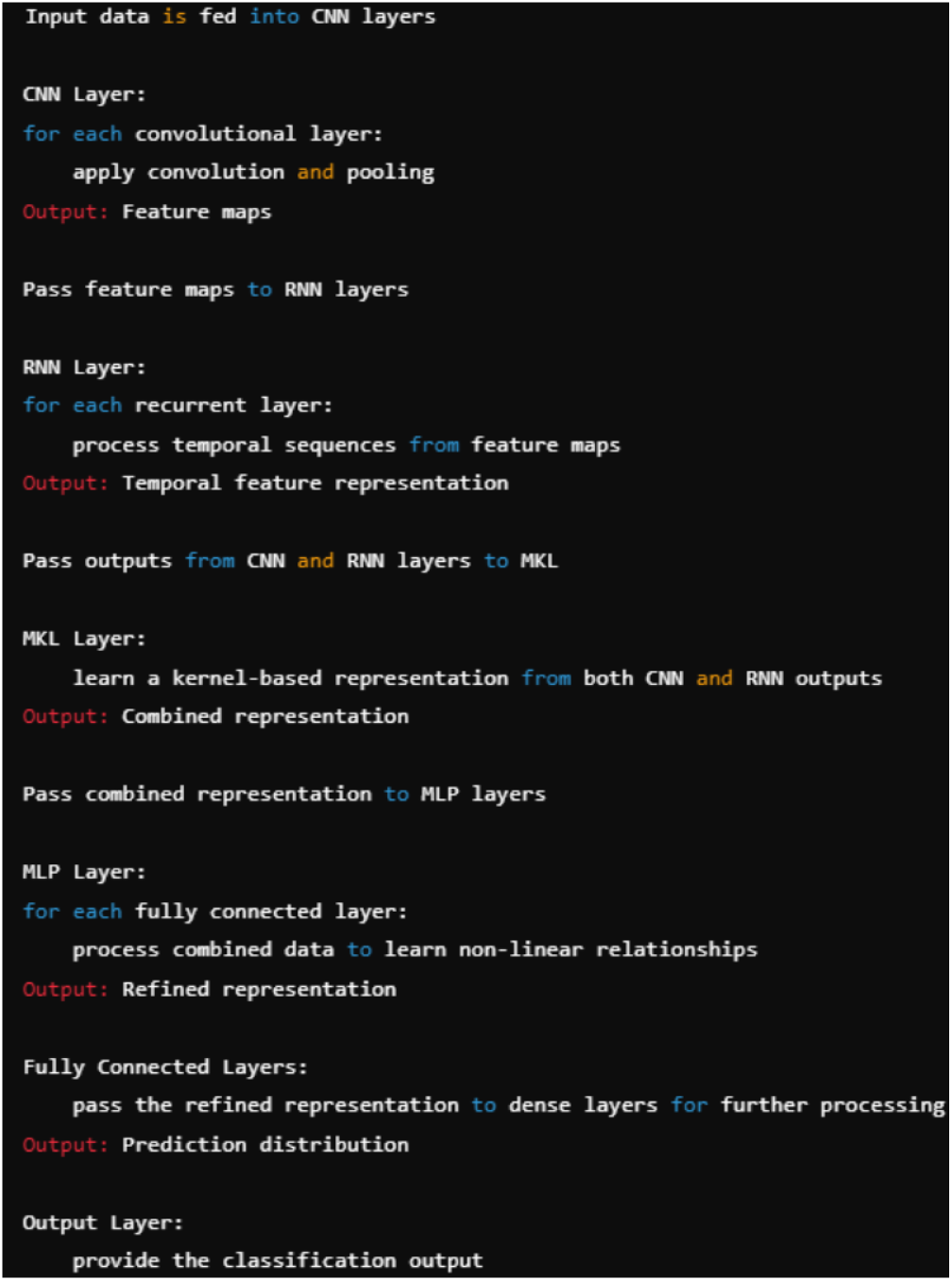

This pseudocode outlines the primary steps in training the hybrid MLP + CNN + RNN + MKL model.

### F. Dataset Information

1. **Source:**

**•** The dataset used in this study was obtained from Figshare, titled *“Voice Samples for Patients with Parkinson’s Disease and Healthy Controls”* (DOI: https://doi.org/10.6084/m9.figshare.23849127).
2. **Participant Details:**

**•** 81 voice recordings (41 from HCs and 40 from PD patients).
**•** Demographic information, including sex ratio, age at collection, Hoehn & Yahr stage of PD, and length of disease, can be found in *Table 1a*.
3. **Ethical** Considerations:
  **•** All data was from a public dataset and were anonymized, thus adhering to the ethical standards of data usage and participant privacy.

## Footnotes

1 https://drive.google.com/drive/folders/15JYAPF7K-xJMGLWXtr36cqBFwXAYW1OG.

2 https://github.com/MatthewShen08/Parkinson-s-Diagnosis-using-XAI

3 https://doi.org/10.6084/m9.figshare.23849127

## References

[1] Little MA et al. “Suitability of dysphonia measurements for telemonitoring of Parkinson’s disease.” In: IEEE Trans Biomed Eng. DOI: 10.1109/TBME.2008.2005954. IEEE. 2009, 56(4):1015–1022.

[2] Tsanas A et al. “Accurate telemonitoring of Parkinson’s disease progression by noninvasive speech tests.” In: IEEE Trans Biomed Eng. DOI: 10.1109/TBME.2009.2036000. IEEE. 2010, 57(4):884–893.

[3] Alhanai T, Au R, and Glass. J. “Detecting Depression with Audio/Text Sequence Modeling of Interviews.” In: DOI: 10.21437/Interspeech.2018-2522. Interspeech. 2018, pp. 1716–1720.

[4] Alissa M et al. “Parkinson’s disease diagnosis using convolutional neural networks and figure-copying tasks.” In: Neural Comput Appl. DOI: 10.1007/s00521-021-06469-7. Springer. 2022, 34(2):1433–1453.

[5] Guo G et al. “Diagnosing Parkinson’s Disease Using Multimodal Physiological Signals.” In: Communications in Computer and Information Science. DOI: 10.1007/978-981-16-1288-69.. Springer, Singapore. 2021, pp. 125–136.

[6] Kumar K and Ghosh. R. “Parkinson’s disease diagnosis using recurrent neural network based deep learning model by analyzing online handwriting.” In: Multimed Tools Appl. DOI: 10.1007/s11042-023-15811-1. Springer. 2023, 83:11687–11715.

[7] Kumar K et al. “A Perspective on Explainable Artificial Intelligence Methods: SHAP and LIME.” In: Adv Intell Syst. DOI: 10.1002/aisy.202400304. Wiley Online Library. 2024.

[8] Dixit S et al. “A comprehensive review on AI-enabled models for Parkinson’s disease diagnosis. Electronics.” In: Electronics. DOI: 10.3390/electronics12040783. MDPI. 2023, 12(4):783.

[9] Santa Cruz BG, Husch A, and Hertel. F. “Machine learning models for diagnosis and prognosis of Parkinson’s disease using Brain Imaging: General overview, main challenges, and future directions.” In: Front Aging Neurosci. DOI: 10.3389/fnagi.2023.1216163. frontiers. 2023, 15:1216163.

[10] Klucken J et al. “Management of Parkinson’s disease 20 years from now: towards digital health pathways.” In: J Parkinsons Dis. DOI: 10.3233/JPD-181519. IOS Press. 2018, 8(s1):S85–S94.

[11] Iyer A et al. “A machine learning method to process voice samples for identification of Parkinson’s disease.” In: Sci Rep. DOI: 10.1038/s41598-023-47568-w. nature. 2023, 13:20615.

[12] Rizzo G et al. “Accuracy of clinical diagnosis of Parkinson disease: A systematic review and meta-analysis.” In: Neurlogy. DOI: 10.1212/WNL.0000000000002350. Neurology Journals. 2016, 86(6):566–576.

[13] Gómez-Rodellar A et al. “Performance of Articulation Kinetic Distributions Vs MFCCs in Parkinson’s Detection from Vowel Utterances Neural Approaches to Dynamics of Signal Exchanges.” In: *Smart Innovation*, Systems and Technologies. DOI: 10.1007/978-981-13-8950-438.. Singapore, Springer. 2019, 151:431–441.

[14] Bouagina S et al. “MFCC-Based Analysis of Vibratory Anomalies in Parkinson’s Disease Detection using Sustained Vowels.” In: IEEE Afro-Mediterranean Conference on Artificial Intelligence. DOI: 10.1109/AM-CAI59331.2023.10431494. IEEE. 2023, pp. 1–5.

[15] Ma A, Lau KK, and Thyagarajan. D. “Radiological correlates of vocal fold bowing as markers of Parkinson’s disease progression: A cross-sectional study utilizing dynamic laryngeal.” In: *CT*. PLoS One. DOI: 10.1371/jour-nal.pone.0258786. Pone. 2021, 16(10).

